# Proteomic profiling of soft tissue sarcomas with SWATH mass spectrometry

**DOI:** 10.1101/2020.06.11.20128355

**Authors:** Martina Milighetti, Lukas Krasny, Alex T.J. Lee, Frank McCarthy, Gabriele Morani, Cornelia Szecsei, Yingtong Chen, Cyril Fisher, Ian Judson, Khin Thway, C.U. Cheang Maggie, Robin L. Jones, Paul H. Huang

**Affiliations:** Division of Molecular Pathology, The Institute of Cancer Research, London, UK; Sarcoma Unit, The Royal Marsden NHS Foundation Trust, London, UK; Division of Clinical Studies, The Institute of Cancer Research, London, UK; Clinical Trials and Statistics Unit, The Institute of Cancer Research, London, UK

**Keywords:** Soft tissue sarcoma, Proteomics, Mass spectrometry, SWATH MS, FFPE, biomarkers

## Abstract

Soft tissue sarcomas (STS) are a group of rare and heterogeneous cancers. While large-scale genomic and epigenomic profiling of STS have been undertaken, proteomic analysis has thus far been limited. Here we utilise sequential window acquisition of all theoretical fragment ion spectra mass spectrometry (SWATH-MS) for proteomic profiling of formalin fixed paraffin embedded (FFPE) specimens from a cohort of STS patients (n=36) across four histological subtypes (leiomyosarcoma, synovial sarcoma, undifferentiated pleomorphic sarcoma and dedifferentiated liposarcoma). We quantified 2951 proteins across all cases and show that there is a significant enrichment of gene sets associated with smooth muscle contraction in leiomyosarcoma, RNA splicing regulation in synovial sarcoma and leukocyte activation in undifferentiated pleomorphic sarcoma. We further identified a subgroup of STS cases (independent of histological subtype) that have a distinct expression profile in a panel of 133 proteins, with worse survival outcomes when compared to the rest of the cohort. Our study highlights the value of comprehensive proteomic characterisation as a means to identify histotype-specific STS profiles that describe key biological pathways of clinical and therapeutic relevance; as well as for discovering new prognostic biomarkers in this group of rare and difficult-to-treat diseases.

## Introduction

Soft tissue sarcomas (STS) are a group of rare and heterogeneous malignancies of mesenchymal origin, comprising more than 100 distinct diagnostic subtypes that are primarily defined by histological characteristics (1). Multiple gene expression-based studies across different histological subtypes have led to a deeper understanding of the oncogenic processes driving STS development and progression as well as the identification of prognostic factors for these cancers (2–5). Whilst current diagnostic evaluation of STS is reliant on histological assessment by specialist sarcoma histopathologists, supplemented by specific molecular tests in selected subtypes, recent large-scale genomic and epigenetic analyses have demonstrated the feasibility of refining the classification system of STS based on intrinsic underlying biology (3, 4). Collectively, these studies highlight the promise of molecular profiling strategies in improving our knowledge of drivers of sarcoma pathogenesis, providing complementary information to aid in the molecular classification of these heterogeneous tumours, identify new therapeutic targets and develop clinically relevant prognostic biomarkers.

While the advent of next generation sequencing (NGS) has accelerated the use of genomic and epigenetic profiling in STS, proteomic analysis in this disease has been limited (6). Comprehensive analysis of the tumour proteome is highly informative as proteins represent the largest class of druggable targets and directly reflect the functional state of biological pathways (7). Proteomic analysis of STS has been performed by The Cancer Genome Atlas (TCGA) consortium in a cohort of 173 flash-frozen sarcoma specimens across six histological subtypes (3). However, this study utilised the reverse phase protein array (RPPA) platform which is limited to the analysis of 192 proteins/phosphoproteins. Other studies that utilise mass spectrometry (MS) have largely been limited to 1-2 subtypes including a recent proteomic analysis of 13 gastrointestinal stromal tumours (GIST) which utilised mass spectrometry (MS) to identify three clinical subgroups (6, 8). To date no MS-based analysis has been conducted across multiple histological subtypes in STS.

In this study, we utilise sequential window acquisition of all theoretical fragment ion spectra (SWATH)-MS to undertake proteomic profiling of FFPE tissue specimens in a cohort of STS patients across four histological subtypes (n=36). We characterise the intrinsic biological pathways associated with specific histotypes and explore the association of proteomic data with patient outcome. This study represents, to our knowledge, the most comprehensive analysis of the proteome in multiple STS subtypes to date and highlights the potential of proteomic profiles as predictors for patient outcome in these rare cancers.

## Results

### Patient characteristics

The cohort is comprised of FFPE tumour material from 36 patients treated at The Royal Marsden Hospital. These specimens were obtained from surgical resections of primary tumours from four of the more common STS subtypes: leiomyosarcoma (LMS) (n=12), synovial sarcoma (SS) (n=7), undifferentiated pleomorphic sarcoma (UPS) (n=10) and dedifferentiated liposarcoma (DDLPS) (n=7). Baseline clinico-pathological characteristics of these patients are summarised in Table 1.

**Table 1:** Clinico-pathological characteristics of STS cohort

|  |  | Overall | LMS | SS | UPS | DDLPS |
| --- | --- | --- | --- | --- | --- | --- |
| **Number of cases** |  | 36 | 12 | 7 | 10 | 7 |
| **Age** |  | 62<br>(28 - 84) | 67<br>(35 - 75) | 49<br>(43 - 77) | 76<br>(28 - 84) | 61<br>(37-78) |
| **Gender** | Female | 25 | 12 | 4 | 5 | 4 |
|  | Male | 11 | 0 | 3 | 5 | 3 |
| **Anatomical site** | Intracavity | 14 | 5 | 2 | 2 | 5 |
|  | Limbs | 12 | 3 | 4 | 4 | 1 |
|  | Trunk/head | 6 | 0 | 1 | 4 | 1 |
|  | Uterus | 4 | 4 | 0 | 0 | 0 |
| **Disease stage** | Localised | 36 | 12 | 7 | 10 | 7 |
|  | Local recurrent | 0 | 0 | 0 | 0 | 0 |
| **Histological grade** | 1 | 0 | 0 | 0 | 0 | 0 |
|  | 2 | 16 | 6 | 4 | 4 | 2 |
|  | 3 | 20 | 6 | 3 | 6 | 5 |
| **Pre-op treatment** | No treatment | 32 | 12 | 3 | 10 | 7 |
|  | Chemotherapy | 1 | 0 | 1 | 0 | 0 |
|  | Chemo & Radiotherapy | 3 | 0 | 3 | 0 | 0 |

### Quantitative analysis of the STS proteome

STS specimens were subjected to protein extraction and SWATH-MS analysis in technical duplicates as outlined in Figure 1. This analysis led to the identification and quantification of 2951 proteins across all samples (Figure 2A and Table S1). Applying t-Distributed Stochastic Neighbour Embedding (t-SNE) on the full dataset, four major groups can be visualised on the 3D-tSNE plot (Figure 2B), which correspond to the four distinct histological subtypes. These results demonstrate that SWATH-MS profiling can reveal intrinsic biology associated with distinct histological subtypes which are characterised by unique proteomic signatures.

**Figure 1.**
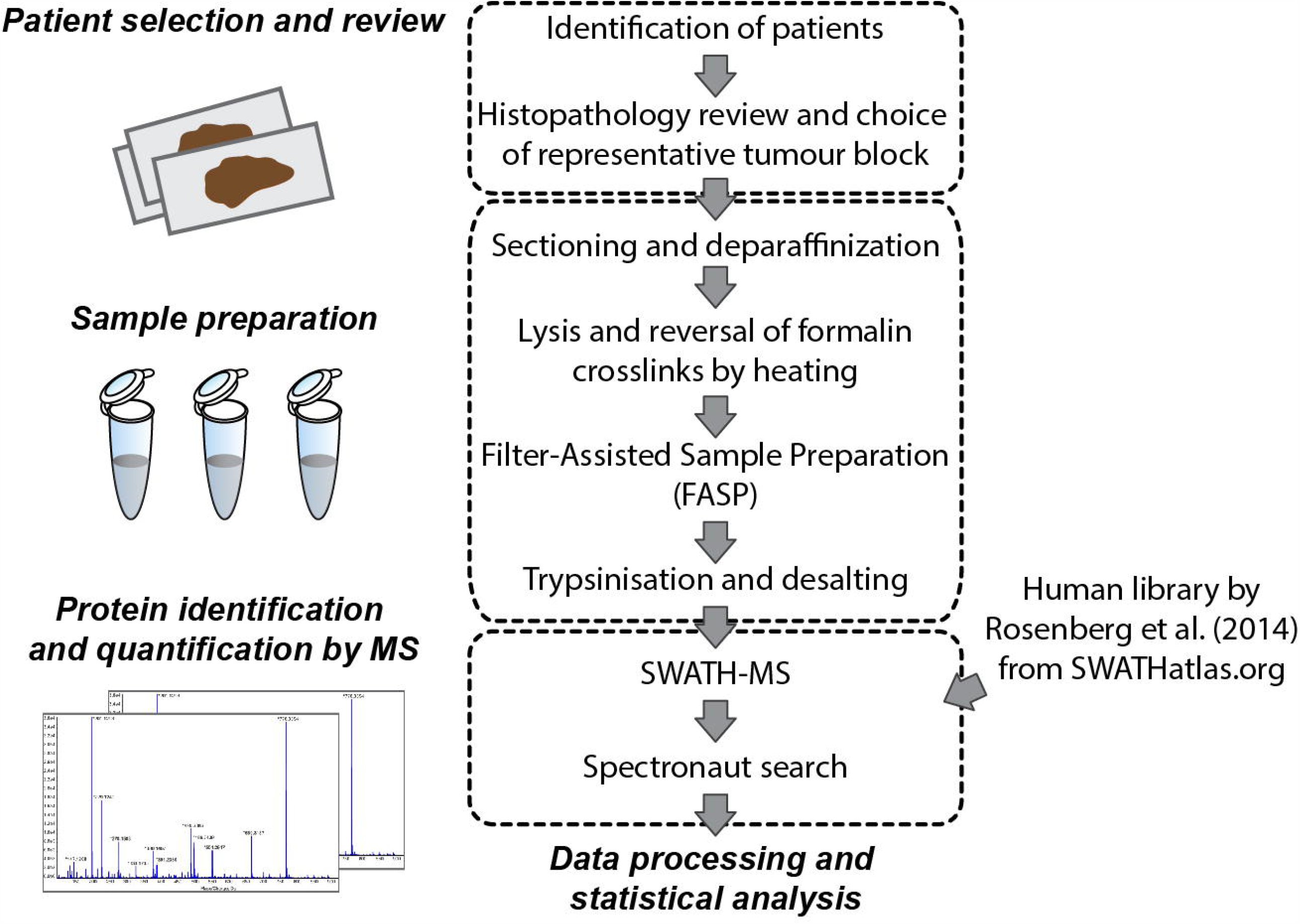
Schematic of the experimental workflow highlighting the key steps that were undertaken for sample selection and preparation as well as SWATH-MS data acquisition and analysis.

**Figure 2.**
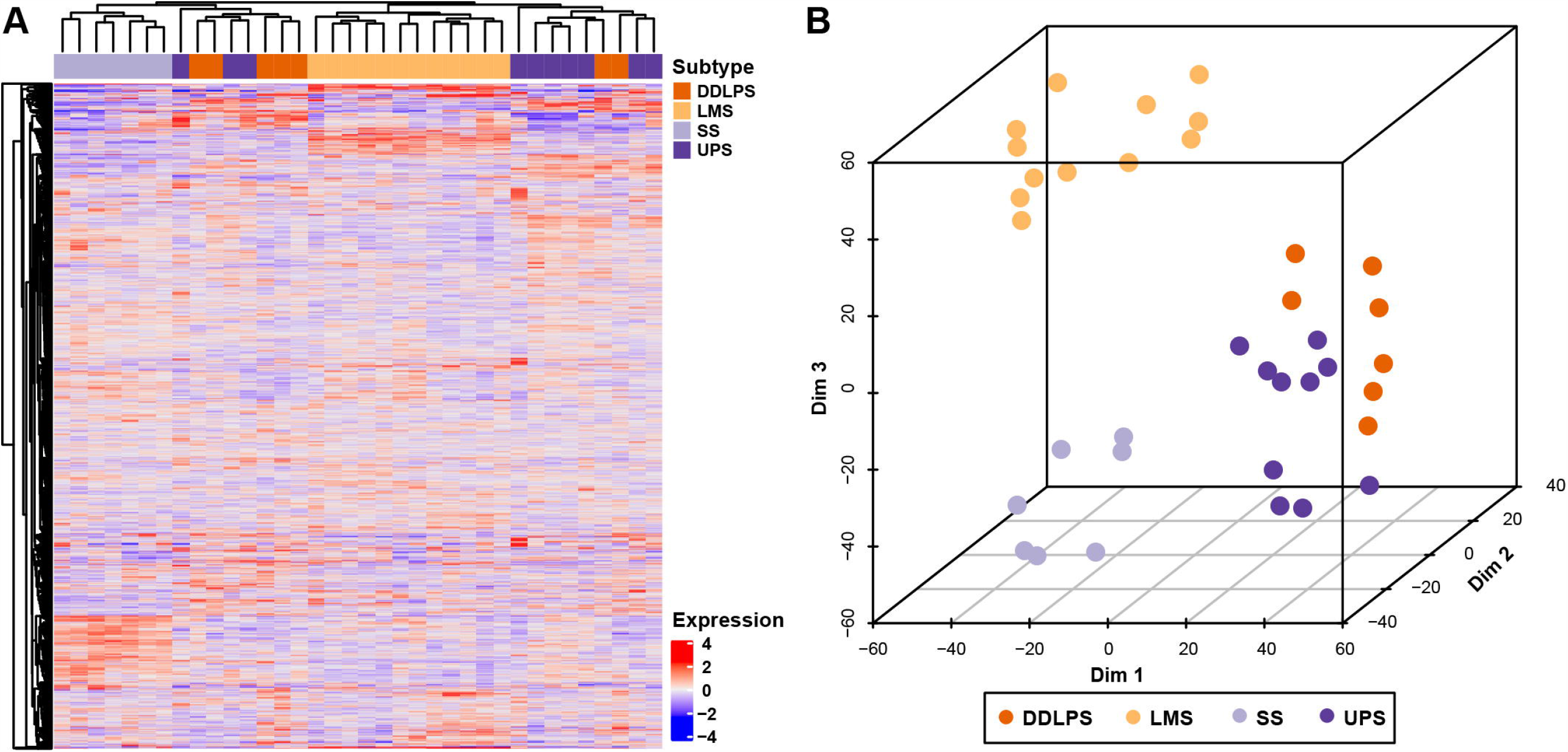
(A) Hierarchical clustering of 2951 proteins across 36 STS cases. The full list of proteins is listed in Table S1. (B) 3D-tSNE plot depicts four distinct groups of STS cases corresponding to the distinct histological subtypes. LMS is leiomyosarcoma, SS is synovial sarcoma, UPS is undifferentiated pleomorphic sarcoma and DDLPS is dedifferentiated liposarcoma.

### Defining biological processes that are enriched in STS histological subtypes

To gain an understanding of the underlying biological pathways in each histological subtype, we undertook gene set enrichment analysis (GSEA) of the full proteomic dataset (9). The top 20 ranked enriched biological processes are shown in Figure 3. Gene sets significantly enriched in LMS versus the rest of the cohort comprise of those involved in muscle development and contraction. This finding is consistent with the smooth muscle lineage of this histological subtype which has also been reported in published gene expression studies of LMS (2, 10). The utility of SWATH-MS data to rediscover known molecular processes highlights the validity of this approach for pathway discovery. In the case of SS, GSEA identifies RNA splicing and processing to be key gene sets significantly enriched in this histological subtype. This result is in line with a previous study which showed that the SS18-SSX fusion in synovial sarcomas binds to the ribonucleoprotein SYT-interacting protein/co-activator activator (SIP/CoAA) which is a key modulator of RNA splicing (11). Meanwhile, gene sets significantly enriched in UPS finds a number of biological processes linked to leukocyte activation and metal ion transport. Notably, Phase II STS clinical trials of immune checkpoint inhibitors have reported evidence of clinical activity in UPS (12, 13), underscoring the value of SWATH-MS in identifying biologically meaningful processes with potential clinical utility. No ontologies were found to be significantly enriched in DDLPS.

**Figure 3.**
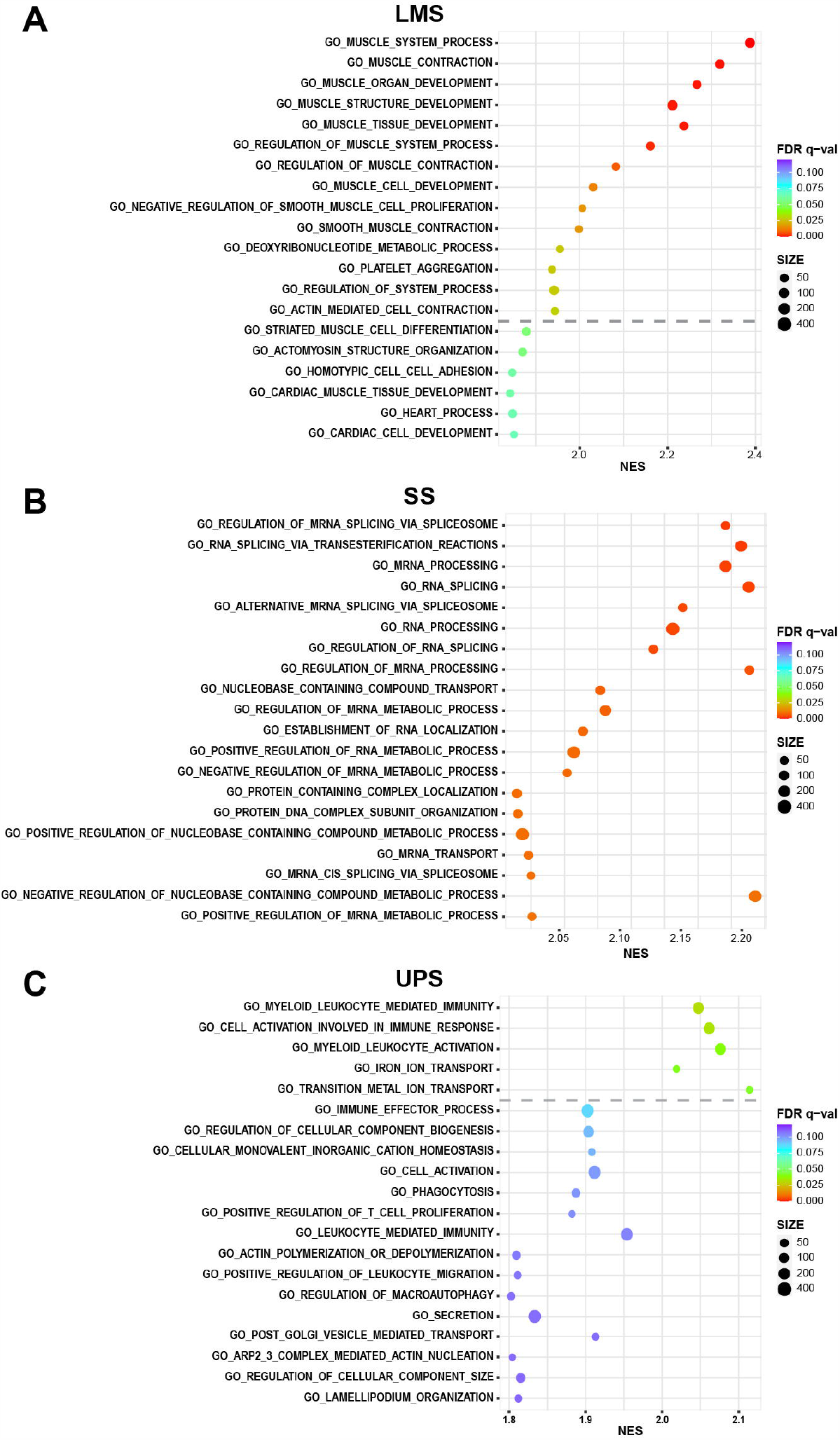
Plot of Gene Set Enrichment Analysis results showing the top ranked 20 positively enriched gene sets for (A) leiomyosarcoma (LMS), (B) synovial sarcoma (SS) and (C) undifferentiated pleomorphic sarcoma (UPS). The dashed line indicates a False Discovery Rate (FDR) = 0.05 threshold. The colour of the circles represent the FDR q-value while the size of the circle indicates the number of proteins within the dataset that is in each gene set. NES is the normalised enrichment score from the GSEA analysis.

### Identification of key protein complexes and signalling networks operating in STS histotypes

Using the multiclass Significance Analysis of Microarray (SAM) method, 277 proteins (FDR <0.1%) were found to be significantly differentially expressed across these four histotypes (Figure S1 and Table S2). Of these proteins, 103 were found to be significantly upregulated only in LMS, and 23 seed proteins were significantly mapped to directly interact with each other to form a zero-order network according to the STRING interactome database (Figure 4A). These 23 seed proteins formed a subnetwork including the core machinery required for the regulation of smooth muscle contractile activity such as the myosin light chains (MYL1, MYL6, MYL9, MYL12B), myosin heavy chains (MYH10, MYH11), tropomyosin alpha chains (TPM1, TPM4), Myosin Light Chain Kinase (MYLK) and Protein Phosphatase 1 Regulatory Subunit 12A or Myosin Phosphatase Target Subunit 1 (PPP1R112A) (Figure 4A). A second protein subnetwork comprising extracellular matrix components (matrisome) and their cognate adhesion receptors and downstream regulatory proteins (adhesome) was also found to be upregulated in LMS. This network includes the integrin receptors (ITGA5, ITGB1, ITGB5), intracellular adhesion signalling proteins (VCL, FLNB, ILK, TNS1) and extracellular matrisome components (HSPB1, HSPG1, NID1, LAMB2, TNC, TGFB1I1) (14, 15) (Figure 4A).

**Figure 4.**
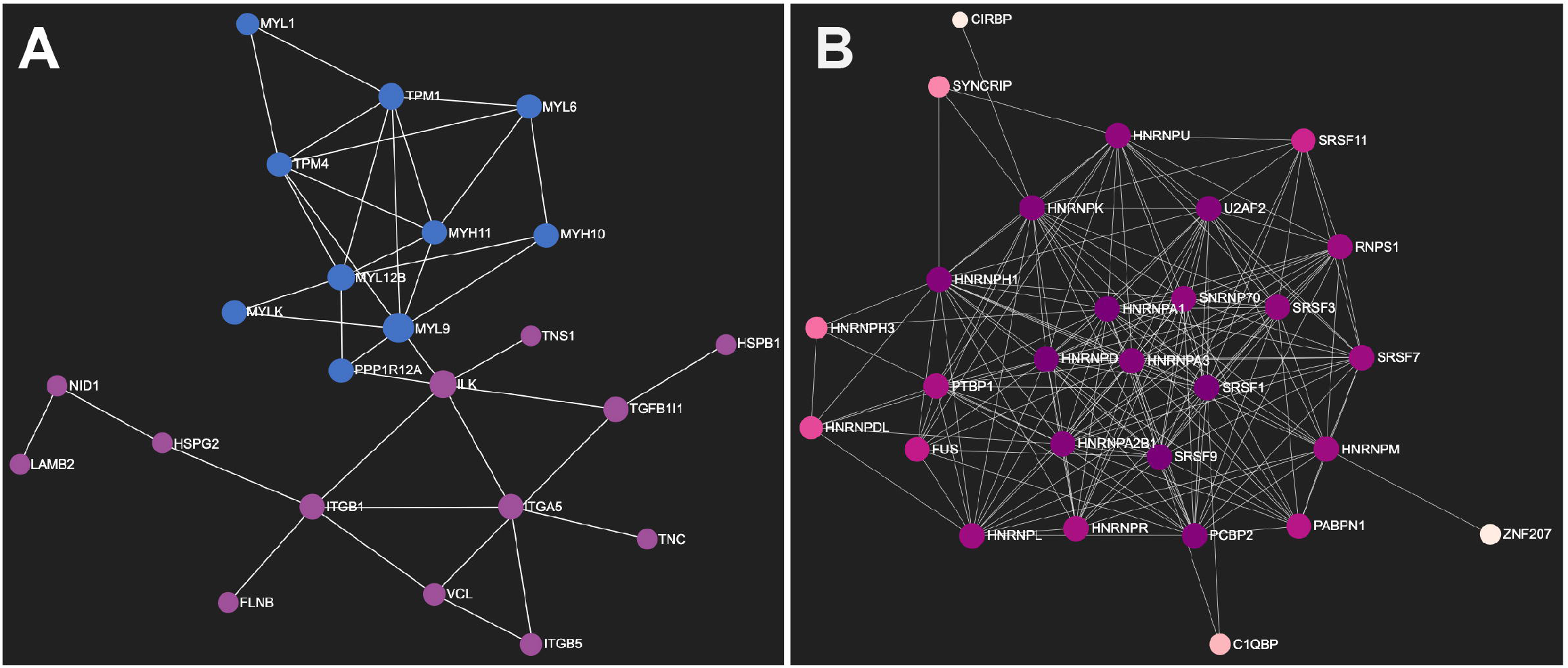
Network diagrams in Force Atlas layouts depicting protein-protein interaction maps for proteins which are significantly upregulated in (A) leiomyosarcoma and (B) synovial sarcoma. In (A), proteins in blue are components involved in the regulation of smooth muscle contraction while those in purple are components of the matrisome and adhesome. In (B), the majority of significantly upregulated proteins in synovial sarcoma are involved in RNA splicing regulation and the different colours indicate the number of interactions between identified proteins.

In SS, 103 proteins were found to be uniquely significantly upregulated; 28 of which were identified as seed proteins that significantly mapped to directly interact with each other to form a zero-order network. Twenty-four of these proteins are key components of mRNA splicing regulation. Cross-referencing these proteins to the SpliceosomeDB of spliceosome components (16) showed that the proteins enriched in SS comprise of the SR protein class (SRSF1, SRSF3, SRSF7, SRSF9, SRSF11) involved in constitutive and alternative pre-mRNA splicing, the heterogeneous ribonucleoprotein particle (hnRNPs) class and proteins involved in the formation of the spliceosome A Complex (HNRNPA1, U2AF2, SNRNP70) (Figure 4B). Twenty-nine proteins were found to be upregulated only in UPS, and 3 seed proteins directly interact with each other, all components of the MHC class 1 complex (HLA-A, HLA-B and B2M). In DDLPS, 13 proteins were uniquely upregulated with no zero-order network identified. Notably, CDK4 is one of these 13 upregulated proteins which is in agreement with the molecular pathology of this disease where CDK4 is amplified in ∼90% of DDLPS; and CDK4/6 inhibitors (palbociclib, ribociclib and abemaciclib) are currently being evaluated in the treatment of this histological subtype (17–19).

### Identification of proteomic profiles that are predictive of STS patient outcome

In order to evaluate if there are proteins within our dataset that are associated with overall survival (OS) in our cohort of patients, univariable Cox regression analysis was performed for each of the 2951 proteins as exploratory analyses, and we selected a total of 133 proteins with p<0.05 (Figure 5A and Table S3). Subsequently, by hierarchical clustering of the 36 cases based on these 133 protein expression values, three subgroups of mixed histological subtypes were identified (Figure 5B). These three groups were associated with significantly differential OS, with Group 2 comprising cases demonstrating the worst survival estimate (Figure 5C, Log-rank p<0.00001). Adjusting for other clinicopathological factors including age, tumour size, grade and histological subtypes, the molecular subgroups remained an independent prognostic factor. After adjustment for other prognostic factors, patients in Group 2 were 81 times more likely to die (Hazard ratio 81; 95% Confidence Interval 6-1085; p=0.0009) compared to patients in Group 1. No statistically significant biological pathways were identified to be enriched in the 133 proteins based on over-representation analysis (20).

**Figure 5.**
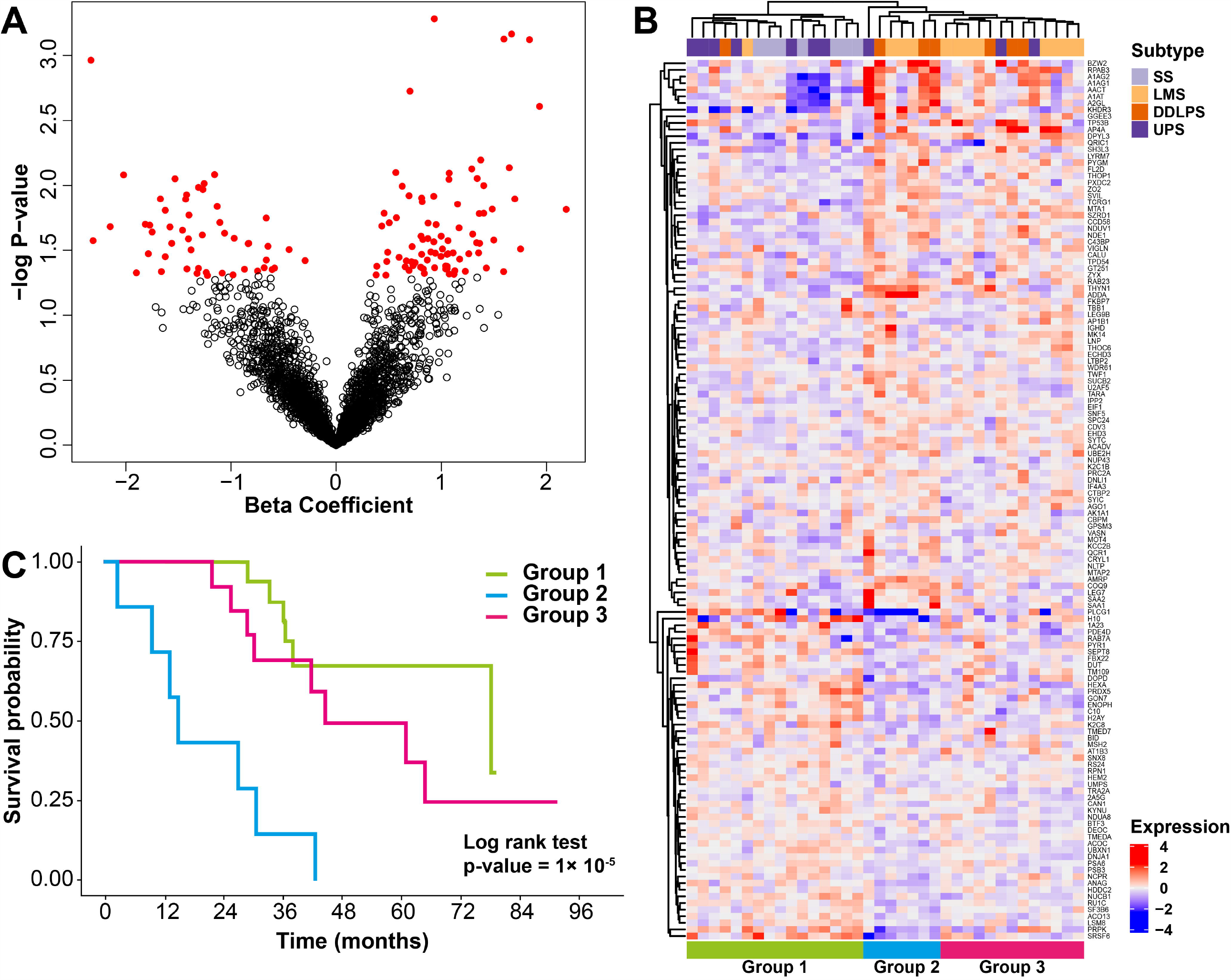
(A) Volcano plot of the beta coefficients from univariable Cox regression analysis for each of the 2951 proteins in the proteomic dataset and their associated –log p-value. Red circles indicates 133 proteins with p<0.05. The full list of proteins is listed in Table S3. (B) Hierarchical clustering of 133 proteins across 36 STS cases identifies 3 subgroups of mixed histological subtypes. (C) The Kaplan Meier curves for overall survival (OS) of the 3 subgroups identified in (B). LMS is leiomyosarcoma, SS is synovial sarcoma, UPS is undifferentiated pleomorphic sarcoma and DDLPS is dedifferentiated liposarcoma.

## Discussion

In this study we have undertaken the most comprehensive proteomic analysis of multiple STS subtypes to date and have demonstrated the utility of SWATH-MS in the following applications: 1. Defining unique proteomics signatures associated with distinct histological subtypes, 2. Identifying, within histological subtypes, biological processes and key protein networks and 3. Defining histotype-independent candidate biomarkers for predicting patient outcomes. Expanded proteomic analysis incorporating larger STS cohorts with an increased number of histological subtypes will enable further refinement of this approach as a means of gaining new insights into the biology and molecular drivers of this rare group of cancers. In particular, the enrichment of an immune response signature in UPS, where immune checkpoint inhibitors have showed some clinical activity (12, 13), suggests that future inclusion of proteomic profiling in cancer immunotherapy trials may lead to new predictive biomarkers that complement currently available approaches (21).

Several transcriptomic studies have led to the development of prognostic biomarkers in sarcoma. These include gene expression panels based on chromosome instability, hypoxia and stromal signatures (5, 22–24). Here we have performed an exploratory analysis which has for the first time identified a panel of proteins that is capable of stratifying a subgroup of patients (Group 2) with worse outcomes when compared with the rest of the cohort. Notably, Group 2 is composed of patients with mixed histological subtypes (3 DDLPS, 3 LMS and 1 UPS), and has a distinct expression profile of 133 proteins. Interestingly, Group 2 shared a subgroup of proteins that were also overexpressed within Group 3 (comprising mainly of DDLPS and LMS), while having low expression in a subset of proteins compared to both Group 1 and 3. These data suggest that these patients may share molecular characteristics that transcend histotypes. Over-representation analysis did not identify any enriched biological pathways in the 133 proteins. It is important to note that this analysis was performed on a small patient cohort treated within a single institution and our findings should be considered hypothesis generating. Further assessment of these 133 proteins in additional validation cohorts is required to confirm any prognostic association.

Together with a recently published SWATH-MS analysis in prostate cancer and diffuse large B-cell lymphoma (25), our study demonstrates the versatility of this label-free MS strategy in the analysis of FFPE specimens, a tissue preservation technique which has historically posed technical challenges for conventional proteomic workflows and analyses (26). By utilising archival material from a tissue bank which is typically an abundant tissue resource, our study describing the proteomic profiling of STS from FFPE specimens opens new opportunities for such proteomic studies in rare cancers without the need for logistically challenging prospective collection of fresh tissue.

In conclusion, we have employed SWATH MS to profile FFPE tissue specimens across four sarcoma subtypes and have identified histotype-specific proteomic profiles that describe key biological pathways as well as discovered a new candidate prognostic biomarker protein panel in STS. We anticipate the future application of this strategy to additional STS subtypes and clinical trial cohorts has the potential to deliver new therapeutic targets and define predictive and prognostic biomarkers in these rare and difficult-to-treat diseases.

## Materials and Methods

### Patients and tumour specimens

Use of archival FFPE tumour samples and linked anonymised patient data was approved by Institutional Review Board as part of the PROSPECTUS study, a Royal Marsden-sponsored non-interventional translational protocol (CCR 4371, REC 16/EE/0213). FFPE tissue from surgically resected primary tumours from four STS subtypes (LMS, SS, UPS and DDLPS) and accompanying annotation of baseline clinicopathological variables were identified and retrieved through retrospective review of departmental database and medical notes at a single specialist cancer centre. In line with standard management approaches, primary LMS, UPS and DDLPS tumours were naïve to any pre-operative therapy, while pre-operative exposure to chemotherapy and/or radiotherapy was varied in SS. The histological diagnosis was confirmed in all cases by experienced soft tissue pathologists (KT, CF). For each tumour, a single FFPE tissue block containing representative viable tumour was selected through review of haematoxylin and eosin (H&E)-stained sections. 20µm sections were cut from each selected tumour block and, where indicated, macrodissected to enrich to >75% viable tumour content. In LPS tumours that contained both well-differentiated and de-differentiated components, slide review and macrodissection ensured the dedifferentiated areas were sampled.

### Protein extraction and sample preparation

20µm tissue sections from each sample were deparaffinised by three washing steps in xylene, rehydrated by washes with decreasing ethanol gradient (100%, 96%, 70%) and then dried in a SpeedVac concentrator (Thermo Scientific). The lysis buffer (0.1M Tris-HCl pH 8.8, 0.50% (w/v) sodium deoxycholate, 0.35% (w/v) sodium lauryl sulphate) was added at a ratio of 200ul/mg of dry tissue. The sample was homogenised using a LabGen700 blender (ColeParmer) with 3x 30s pulses and sonicated on ice for 10 min, then heated at 95^°^C for 1 h to reverse formalin crosslinks. Lysis was carried out by shaking at 750rpm at 80^°^C for 2 h. The sample was then centrifuged for 15min at 4°C at 14,000rpm and the supernatant collected. Protein concentration in the homogenate was measured by bicinchoninic acid (BCA) assay (Pierce) The extracted protein sample was digested using the Filter-Aided Sample Preparation (FASP) protocol as previously described in (27). Briefly, each sample was placed into an Amicon-Ultra 4 (Merck) centrifugal filter unit and detergents were removed by several washes with 8M urea. The concentrated sample was then transferred to Amicon-Ultra 0.5 (Merck) filters to be reduced with 10mM dithiothreitol (DTT) and alkylated with 55mM iodoacetamide (IAA). The sample was washed with 100mM ammonium bicarbonate (ABC) and digested with trypsin overnight (Promega, trypsin to starting protein ratio 1:100 µg). Peptides were collected by two successive centrifugations with 100mM ABC and desalted on C18 SepPak columns (Waters). The desalted peptide samples were then dried in a SpeedVac concentrator and stored at −80°C.

### SWATH-MS data acquisition and processing

Samples were resuspended in a buffer of 2% ACN/ 0.1% formic acid, spiked with iRT calibration mix (Biognosys AG) and analysed on an Agilent 1260 HPLC system (Agilent Technologies) coupled to a TripleTOF 5600+ mass spectrometer with NanoSource III (AB SCIEX). 1 μg of peptides for each sample was loaded onto a ZORBAX C18 (Agilent Technologies) trap column and separated on a 75 μm×15 cm long analytical column with an integrated manually pulled tip packed with Reprosil Pur C18AQ beads (3 μm, 120 Å particles, Dr. Maisch). A linear gradient of 2–40% of Buffer B (98% ACN, 0.1% FA) in 120 min and a flow rate of 250 nl/min was used. Each sample was analyzed in 2 technical replicates. Full profile MS scans were acquired in the mass range of *m/*z 340-1400 in positive ion mode. 8 data points per elution peak were set up for calculation of 60 precursor isolation windows with a fixed size of 13 Da across the mass range of m/z 380–1100 with 1 Da overlap. MS/MS scans were acquired in the mass range of *m/z* 100-1500. Maximum filling time for MS scans was 250 ms and for MS/MS scans 100 ms, resulting in a cycle time of 3.1 s. SWATH spectra were analysed using Spectronaut 11 (Biognosys AG) against a published human library (28). FDR was restricted to 1% and only the top 6 peptides were used for quantification of a protein. Peak area of 2 to 6 fragment ions was used for peptide quantification, and the mean value of the peptides was used to quantify proteins. 2 peptides were set as minimum requirement for inclusion of a protein in the analysis.

### Data processing and statistical methods

Data were log2 transformed, quantile normalised at sample level, followed by feature level (protein) centering across the samples to remove technical bias such as batch effect. The proteomics data was visualised using 3D-t-Distributed Stochastic Neighbour Embedding. To assess similarity of protein expression profiles, pairwise Pearson correlations between all samples were visualized using the ‘ComplexHeatmap’ R package (29). Dendrograms were created using complete linkage clustering and the Euclidean distances between the resulting correlations. Differential expression analysis was performed using Significant Analysis of Microarray upon a false discovery rate (FDR) less than 0.1% (30). Gene Set Enrichment Analysis (GSEA) version 4.0.1 was used to identify gene sets from the Hallmark database (v7.0, ‘c5.bp.v7.0.symbols.gmt’) that were significantly enriched between each subtype and rest of the groups respectively. Protein-Protein Interaction (PPI) network analysis and visualisation was performed using NetworkAnalyst 3.0 pipeline against the STRING interactome database, with settings to confidence score >900 and experimental evidence required (31–33). Univariable survival analysis of overall survival was estimated by Kaplan-Meier curve and Log-rank test for significance. Event was defined as death. Time was defined as surgery to death or last follow-up. Univariable and multivariable Cox proportional hazards analyses were used to assess the prognostic significance of the different variables. Over-representation analysis were performed on the 133 identified proteins using PANTHER v14 (20).

## Data Availability

The MS proteomics data have been deposited to the ProteomeXchange Consortium via the PRIDE partner repository with the dataset identifier PXD019719

## Availability of data and materials

The MS proteomics data have been deposited to the ProteomeXchange Consortium via the PRIDE (34) partner repository with the dataset identifier PXD019719

## Acknowledgements

We thank Chris Wilding, Nafia Guljar and Jess Burns for assistance in the curation of clinical data and specimens.

## Figure Legends

**Figure S1**. (A) Hierarchical clustering of 277 proteins that were found to be significantly differentially expressed across the four histotypes by multiclass Significance Analysis of Microarray (SAM) method (FDR <0.1%) across 36 STS cases. The full list of proteins is listed in Table S2. LMS is leiomyosarcoma, SS is synovial sarcoma, UPS is undifferentiated pleomorphic sarcoma and DDLPS is dedifferentiated liposarcoma.

